# Using Machine Learning to Elucidate the Spatial and Genetic Complexity of the Ascending Aorta

**DOI:** 10.1101/2021.11.01.21265701

**Authors:** Mahan Nekoui, James P. Pirruccello, Paolo Di Achille, Seung Hoan Choi, Samuel N. Friedman, Victor Nauffal, Kenney Ng, Puneet Batra, Jennifer E. Ho, Anthony A. Philippakis, Steven A. Lubitz, Mark E. Lindsay, Patrick T. Ellinor

## Abstract

**Background:** The left ventricular outflow tract (LVOT) and ascending aorta are spatially complex, with distinct pathologies and embryologic origins. Prior work examined genetics of thoracic aortic diameter in a single plane. We sought to elucidate the genetic basis for the diameter of the LVOT, the aortic root, and the ascending aorta.

**Methods:** We used deep learning to analyze 2.3 million cardiac magnetic resonance images from 43,317 UK Biobank participants. We computed the diameters of the LVOT, the aortic root, and at six locations in the ascending aorta. For each diameter, we conducted a genome-wide association study and generated a polygenic score. Finally, we investigated associations between these polygenic scores and disease incidence.

**Results:** 79 loci were significantly associated with at least one diameter. Of these, 35 were novel, and a majority were associated with one or two diameters. A polygenic score of aortic diameter approximately 13mm from the sinotubular junction most strongly predicted thoracic aortic aneurysm in UK Biobank participants (n=427,016; HR=1.42 per standard deviation; CI=1.34-1.50, P=6.67×10^−21^). A polygenic score predicting a smaller aortic root was predictive of aortic stenosis (n=426,502; HR=1.08 per standard deviation; CI=1.03-1.12, P=5×10^−6^).

**Conclusions:** We detected distinct common genetic loci underpinning the diameters of the LVOT, the aortic root, and at several segments in the ascending aorta. We spatially defined a region of aorta whose genetics may be most relevant to predicting thoracic aortic aneurysm. We further described a genetic signature that may predispose to aortic stenosis. Understanding the genetic contributions to the diameter of the proximal aorta may enable identification of individuals at risk for life-threatening aortic disease and facilitate prioritization of therapeutic targets.

---

The ascending thoracic aorta is a developmentally complex organ arising from the septation of the truncus arteriosus and the bulbus cordis in the later stages of cardiogenesis.^1^ Contributions from two separate germ layers are required for proper aortic morphogenesis, and abnormalities of cardiac ventricular formation or semilunar valve development can influence ascending aortic size and morphology. The phenotypic range in aortic size is wide, from pulmonary stenotic disorders such as tetralogy of Fallot associated with large aortas to congenital aortic valve stenosis and hypoplastic left heart syndrome which can result in an ascending aorta no larger than necessary to transport coronary blood flow.^2,3^ Beyond developmental influences, deficits in aortic homeostasis occur over the lifespan under pathogenic processes such as atherosclerosis or hypertension and can result in aortic growth and aneurysm. Ascending thoracic aneurysm develops asymptomatically, but is associated with a risk of aortic dissection, an important cause of sudden cardiac death. Approximately 50% of patients with a type A dissection of the ascending aorta die prior to arrival at a hospital.^4^ Therefore, understanding the epidemiological and genetic contributions to ascending aortic risk may be important to the development of preventative strategies to avoid sudden cardiac death.^5^ Clinical studies have noted that aneurysms of the ascending aorta occur in younger patients than descending thoracic or abdominal aortic aneurysms, often associated with pathogenic genetic predisposition.^6–8^ Genetic variants in several genes have been associated with ascending aortic aneurysms, including highly penetrant Mendelian loci identified in family studies and common variants identified via GWAS.^9–12^ Prior studies have also described the clinical actionability of genes previously associated with thoracic aortic disease.^5^ However, the majority of patients with thoracic aortic aneurysm do not carry a variant in a known gene.

In prior work, we demonstrated the use of a deep learning architecture to evaluate the dimensions of the thoracic aorta in 4.6 million cardiac magnetic resonance (CMR) images from the UK Biobank.^13^ Using short axis images we conducted genome-wide association studies (GWAS) in up to 39,688 individuals and identified 82 loci associated with ascending thoracic aortic diameter and 47 loci with descending thoracic aortic diameter.

These results contributed to an understanding of the genetic basis of the diameter of the thoracic aorta, potentially facilitating future identification of asymptomatic individuals at risk for aneurysm or dissection using clinical and genetic data. The short axis view used in our prior work limited measurement of the ascending and descending aorta diameters to a single location. As the proximal aorta is known to be spatially complex, consisting of unique anatomical subregions with distinct embryologic origins, we sought to study the structure in greater detail.^14–17^ Therefore we undertook a fine-grained evaluation of ascending aortic dimensions using a deep learning architecture to better understand genetic risk and association with ascending aortic diseases.

## Results

Pathogenic ascending aortic morphology is variable but occurs in reproducible patterns including enlargement of the aortic root, enlargement of the ascending aorta, or combinations of both (**Figure 1A**). Aortic valvular malformations can be associated with smaller or larger aortic diameters. We hypothesized that the diameter of the contiguous tract spanning approximately from the aortic annulus to the aortic arch are complex traits with contributions from common genetic variants. Because this segment is composed of structures known to have distinct biological origins and clinical risk factors for disease, we chose to quantify LVOT diameter, aortic root diameter, and ascending aorta diameter separately. We further hypothesized that within the ascending aorta subsegment, serial calibers spanning approximately from the aortic root to the arch would contain distinct genetic underpinnings with varying degrees of correlation to disease. Therefore, we chose to measure up to six diameters of aorta per participant, depending on the length of aorta captured in the image (see **Methods**).

**Figure 1:**
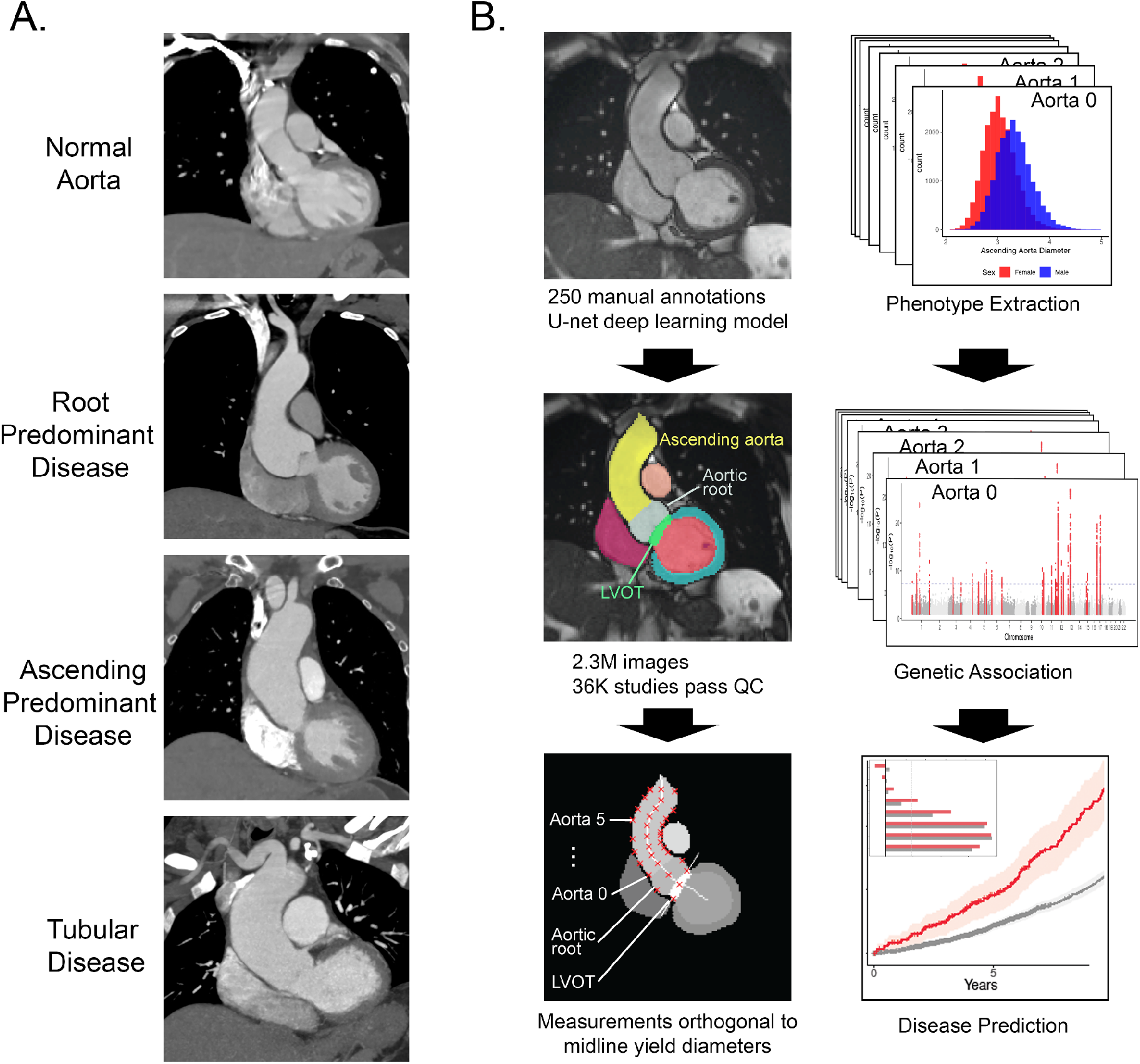
Study overview

### Semantic segmentation with deep learning

First, 250 CMR still-frame images capturing the LVOT, aortic root, and ascending aorta in long axis were randomly selected from the UK Biobank. The images were then manually annotated by one operator (MN) under the supervision of a cardiologist (JPP). This annotation is known as semantic segmentation: the task of identifying and labeling all pixels that comprise an object in an image.

We then used those annotations to train a deep learning model to perform the same semantic segmentation task. We chose a U-Net architecture ^18,19^. As a form of transfer learning, this model’s encoder had been pre-trained on ImageNet, which is a natural-image classification dataset.^20^ Therefore, instead of starting with random weights, the model was initialized with weights that are helpful for processing images, reducing the amount of manual annotation and model training necessary to achieve good model performance.^18,21^

### Phenotype extraction

Having identified which pixels of the given images represented the structures of interest, we then sought to extract the diameter of the LVOT, the diameter of the aortic root, and serial diameters of the ascending aorta. Using classical image processing algorithms, we first computed the centerline of the contiguous structure comprising the left ventricle, LVOT, aortic root, and ascending aorta.^22,23^ We then drew lines orthogonal to the midline at the LVOT, aortic root, and up to six points of the aorta. Serial measurements of the aorta were spaced relative to each participant’s height, and the intervals were chosen to be approximately 1 cm apart for a participant of average height. Finally, we took note of the intersection points of these orthogonal lines and the boundaries of their respective structures of interest. Each normal yielded two intersection points; the Euclidean distance between these two points was taken to represent a diameter of interest. For brevity, we refer to these eight extracted diameters as follows, in order of most proximal to most distal: LVOT, Aortic root, and Aorta 0 through 5. (See **Figure 1B, Supplemental Table 5, Methods** for further illustration of diameter locations).

After quality control, phenotypes were available for up to 33,870 participants. Because the length of aorta captured in the MRI frame is variable, fewer phenotypes were available for more distal measurements of the ascending aorta (**Supplemental Table 3)**. We inspected the distributions of the phenotypes and deemed them to be biologically plausible (**Supplemental Figure 2**).

We sought to validate our diameters by investigating their correlations to prevalent disease (**Supplemental Table 6**). Only 12 participants had both phenotypic data available and diagnoses corresponding to thoracic aortic aneurysm. Nonetheless, all diameters were positively correlated with presence of thoracic aortic aneurysm when adjusted for clinical covariates (see **Methods**). Effect size and significance was greatest for diameters Aorta 1-3.

### GWAS and rare variant association tests of LVOT, aortic root, and ascending aorta diameters

We next sought to define the genetic basis for variation of size of the LVOT, aortic root, and proximal ascending aorta. In total, 33,870 participants had data that passed quality control and contributed to genetic analysis of at least one phenotype (**Table 1**).

**Table 1:** GWAS participant characteristics Demographic information is shown for UK Biobank participants with genetic and cardiac MRI data that passed quality control as detailed in the sample flow diagram in **Supplementary Figure 1**. For count data, values shown are N (%). For quantitative data, values shown are mean (SD).

|  | **Female**<br>**(N=18403)** | **Male**<br>**(N=15467)** | **All**<br>**(N=33870)** |
| --- | --- | --- | --- |
| Age at time of MRI | 63.9 (7.5) | 64.9 (7.8) | 64.3 (7.7) |
| BMI (kg/m<sup>2</sup>) | 25.7 (4.4) | 26.7 (3.7) | 26.2 (4.1) |
| Height (cm) | 163 (6) | 176 (7) | 169 (9) |
| Weight (kg) | 68.1 (12.2) | 82.7 (12.6) | 74.8 (14.4) |
| Systolic Blood Pressure (mmHg) | 136 (19) | 142 (17) | 139 (19) |
| Diastolic Blood Pressure (mmHg) | 76.9 (9.9) | 80.9 (9.7) | 78.7 (10.0) |
| LVOT diameter (mm) | 23.4 (2.6) | 26.4 (3.1) | 24.8 (3.2) |
| Aortic root diameter (mm) | 28.3 (3.6) | 32.4 (4.0) | 30.2 (4.3) |
| Ascending aorta diameter 0 (mm) | 24.7 (3.4) | 27.5 (3.8) | 26.0 (3.8) |
| Ascending aorta diameter 1 (mm) | 26.6 (3.2) | 29.1 (3.5) | 27.7 (3.5) |
| Ascending aorta diameter 2 (mm) | 27.9 (3.3) | 30.4 (3.5) | 29.0 (3.6) |
| Ascending aorta diameter 3 (mm) | 29.0 (3.8) | 31.5 (4.1) | 30.1 (4.1) |
| Ascending aorta diameter 4 (mm) | 28.7 (4.2) | 30.9 (4.2) | 29.7 (4.3) |
| Ascending aorta diameter 5 (mm) | 27.8 (3.9) | 29.9 (4.0) | 28.8 (4.1) |

We confirmed that all extracted phenotypes were heritable traits. The genome-wide single nucleotide polymorphism (SNP) heritability of the size of the LVOT was 22% (95% CI 18-25%); that of the aortic root was 36% (95% CI 32-39%); and those of the ascending aorta ranged from a minimum of 37% (Aorta 0, 95% CI 33-40%) to a maximum of 49% (Aorta 2, 95% CI 45-52%). We further verified that genetic correlation between the extracted traits was generally greater for traits in closer anatomic proximity (**Supplemental Figure 3**).

We then conducted genome-wide association studies (GWAS) of these eight traits, testing 11.2 million genotyped and imputed SNPs with minor allele frequency (MAF) > 0.005 (Supplemental table 4). Using a commonly used genome-wide significance threshold (P < 5×10^−8^), we identified 66 independent loci associated with one or more diameters of the ascending aorta. Of these, 15 were novel as defined by lack of detection in the only other study (to our knowledge) that investigated the genetics of ascending aorta diameter at a comparable anatomic location (see **Methods**) ^13^. In the aortic root, we identified 28 independent genome-wide significant loci. Of these, 17 were novel as defined by lack of detection in the only other study that investigated the genetics of aortic root diameter at a comparable anatomic location. In the LVOT, we identified 6 independent genome-wide significant loci.^24^ (**Figure 2, Table 2**). To our knowledge, no previous GWAS has investigated LVOT diameter. No lead SNPs deviated from Hardy-Weinberg equilibrium (HWE) at the commonly used threshold P < 1×10^−6^.

**Figure 2:**
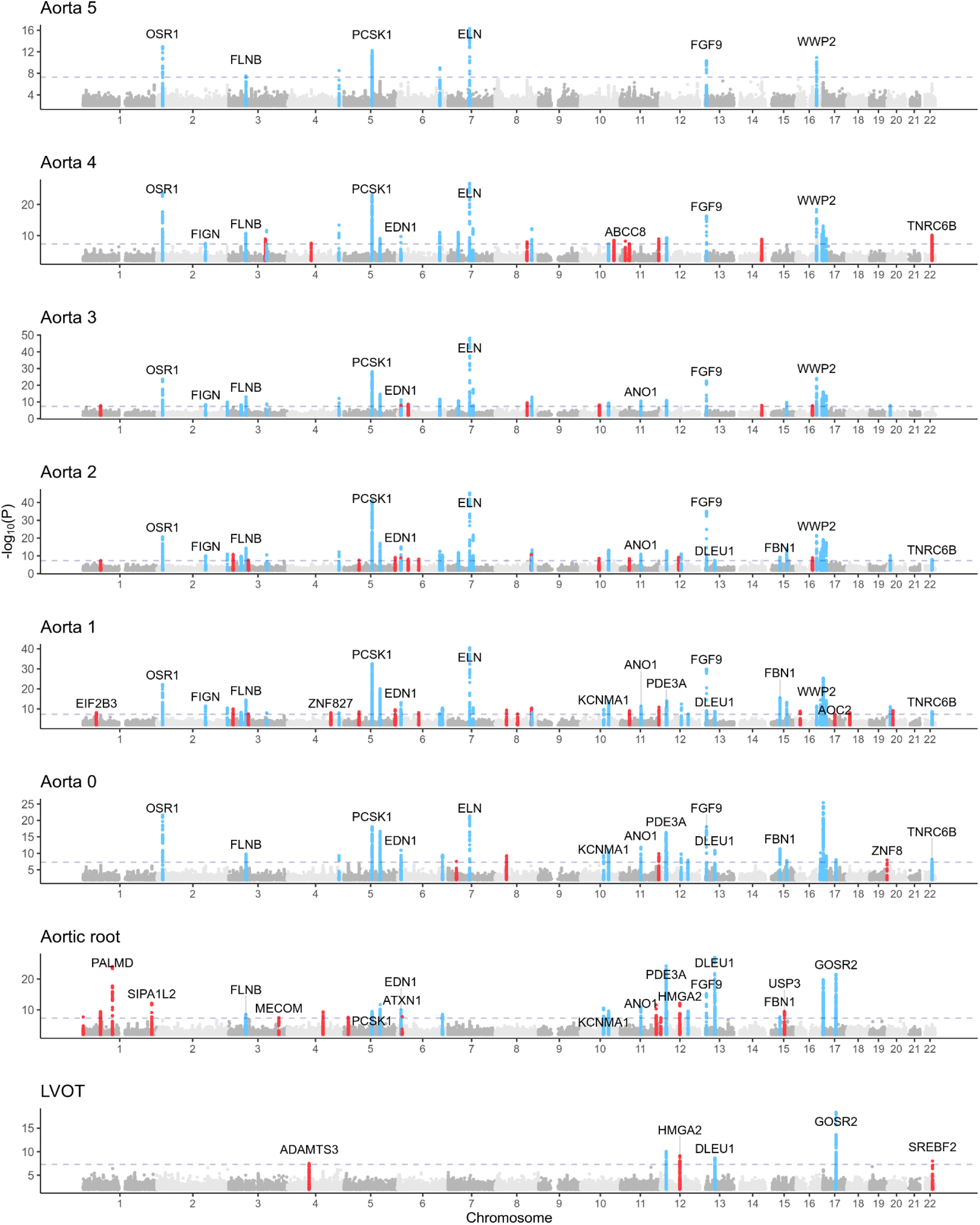
Genetic associations with LVOT, aortic root, and ascending aorta diameters Loci with P < 5×10^−8^ are shown in blue (if also associated at genome-wide significance with two or more anatomically contiguous traits) or red (if associated at genome-wide significance with up to one anatomically contiguous trait. For clarity, a selected subset of nearest genes of loci with P < 5×10^−8^ are overlaid.

**Table 2:** Loci not previously identified in aortic root or aortic diameter GWAS Novel lead SNPs from GWASs for the eight traits of interest. SNP = the rsID of the variant, where available; for variants that are not in dbSNP, this column displays [chromosome] : [genomic position] _ [effect allele _ non-effect allele]. CHR = chromosome. BP = genomic position, keyed to GRCh37.

| Trait | SNP | CHR | BP | Effect/Non-effect allele | Closest gene | P |
| --- | --- | --- | --- | --- | --- | --- |
| Aorta 4 | rs34266187 | 3 | 123975301 | G/A | KALRN | 1.25E-09 |
| Aorta 2 | rs540082300 | 5 | 51196504 | C/CT | ISL1 | 2.83E-08 |
| Aorta 2 | 5:173276788_AT_A | 5 | 173276788 | AT/A | CPEB4 | 8.21E-10 |
| Aorta 2 | rs12195791 | 6 | 72205132 | G/A | OGFRL1 | 7.64E-09 |
| Aorta 2 | rs2137537 | 12 | 71113087 | T/C | PTPRR | 8.19E-12 |
| Aorta 1 | rs747347287 | 1 | 45418228 | AT/A | EIF2B3 | 7.73E-09 |
| Aorta 1 | rs1979974 | 4 | 146800815 | A/G | ZNF827 | 9.39E-09 |
| Aorta 1 | rs13158444 | 5 | 51201361 | T/C | ISL1 | 2.95E-09 |
| Aorta 1 | rs62376928 | 5 | 173288146 | T/C | CPEB4 | 2.67E-10 |
| Aorta 1 | rs143917622 | 6 | 72205158 | A/AT | OGFRL1 | 7.80E-09 |
| Aorta 1 | rs112621658 | 8 | 38774696 | C/A | PLEKHA2 | 4.70E-10 |
| Aorta 1 | rs40430 | 10 | 79179345 | A/T | KCNMA1 | 1.88E-10 |
| Aorta 1 | rs10743356 | 12 | 20235039 | A/G | PDE3A | 2.92E-11 |
| Aorta 1 | rs7304603 | 12 | 71114400 | T/C | PTPRR | 3.78E-13 |
| Aorta 1 | 16:14505488_CT_C | 16 | 14505488 | CT/C | PARN | 1.36E-09 |
| Aorta 1 | rs35296742 | 17 | 40999303 | G/A | AOC2 | 4.41E-08 |
| Aorta 0 | rs917275 | 7 | 28658522 | A/G | CREB5 | 2.91E-08 |
| Aorta 0 | rs112621658 | 8 | 38774696 | C/A | PLEKHA2 | 5.52E-10 |
| Aorta 0 | rs40430 | 10 | 79179345 | A/T | KCNMA1 | 4.34E-10 |
| Aorta 0 | rs10841441 | 12 | 20210632 | C/T | PDE3A | 4.54E-17 |
| Aorta 0 | rs2137537 | 12 | 71113087 | T/C | PTPRR | 1.57E-10 |
| Aorta 0 | rs78033733 | 17 | 45290078 | T/G | MYL4 | 8.66E-09 |
| Aorta 0 | rs57785785 | 19 | 58815158 | T/A | ZNF8 | 1.08E-08 |
| Aortic root | rs76947392 | 1 | 1256608 | G/A | CPSF3L | 2.19E-08 |
| Aortic root | rs1285677 | 1 | 232712355 | A/C | SIPA1L2 | 7.20E-13 |
| Aortic root | rs2686630 | 3 | 58091861 | G/C | FLNB | 3.20E-09 |
| Aortic root | rs73030346 | 3 | 169317856 | C/T | MECOM | 3.54E-08 |
| Aortic root | rs13134800 | 4 | 120900282 | T/C | MAD2L1 | 4.61E-10 |
| Aortic root | 5:15005465_CTCTT_C | 5 | 15005465 | CTCTT/C | ANKH | 2.68E-08 |
| Aortic root | rs4264961 | 5 | 95617018 | C/T | PCSK1 | 3.15E-10 |
| Aortic root | rs1630736 | 6 | 12295987 | C/T | EDN1 | 9.69E-11 |
| Aortic root | rs80036911 | 6 | 16472975 | C/T | ATXN1 | 1.63E-08 |
| Aortic root | rs7754762 | 6 | 152311537 | T/A | ESR1 | 3.06E-09 |
| Aortic root | rs28735 | 10 | 79178044 | G/C | KCNMA1 | 2.80E-11 |
| Aortic root | rs1977289 | 10 | 96301907 | T/C | HELLS | 2.63E-10 |
| Aortic root | rs12280388 | 11 | 121670712 | T/C | SORL1 | 2.48E-12 |
| Aortic root | rs4296081 | 12 | 22005781 | A/G | ABCC9 | 1.94E-10 |
| Aortic root | rs903175 | 12 | 94158719 | G/A | CRADD | 2.73E-10 |
| Aortic root | rs12866004 | 13 | 22865917 | T/A | FGF9 | 5.20E-16 |
| Aortic root | rs17352842 | 15 | 48694211 | C/T | FBN1 | 1.93E-08 |
| LVOT | rs35631249 | 4 | 73443865 | C/A | ADAMTS3 | 3.28E-08 |
| LVOT | rs7139226 | 12 | 20180749 | C/T | AEBP2 | 9.36E-11 |
| LVOT | rs10400419 | 12 | 66389968 | T/C | HMGA2 | 7.77E-10 |
| LVOT | rs2740516 | 13 | 50761619 | G/A | DLEU1 | 2.27E-09 |
| LVOT | rs17677363 | 17 | 45036112 | A/T | GOSR2 | 4.11E-19 |
| LVOT | rs62240962 | 22 | 42259524 | C/T | SREBF2 | 9.75E-09 |

In total, 79 independent loci displayed genome-wide significant association with one or more of the eight extracted diameters. Of these, a majority (41) were significantly associated with only one or two diameters, and only 26 were significantly associated with four or more diameters. Hierarchical clustering of the absolute effect sizes of all significant SNPs provided a visual representation of the observation that the SNPs significantly associated with a given diameter were largely distinct from those significantly associated with other diameters (**Figure 3A**). To investigate this observation with regard to independently associated loci, we binned the genes closest to the loci significantly associated with proximal diameters (LVOT and Aortic root), middle diameters (Aorta 0-2), and distal diameters (Aorta 3-5). We discovered a set of loci independently associated with all three sets (proximal, middle, and distal) as well as associations shared between adjacent pairs of sets (proximal and middle, middle and distal). Notably, no loci were significantly associated with the proximal and distal sets without concurrent association with the middle set (**Figure 3B**).

**Figure 3:**
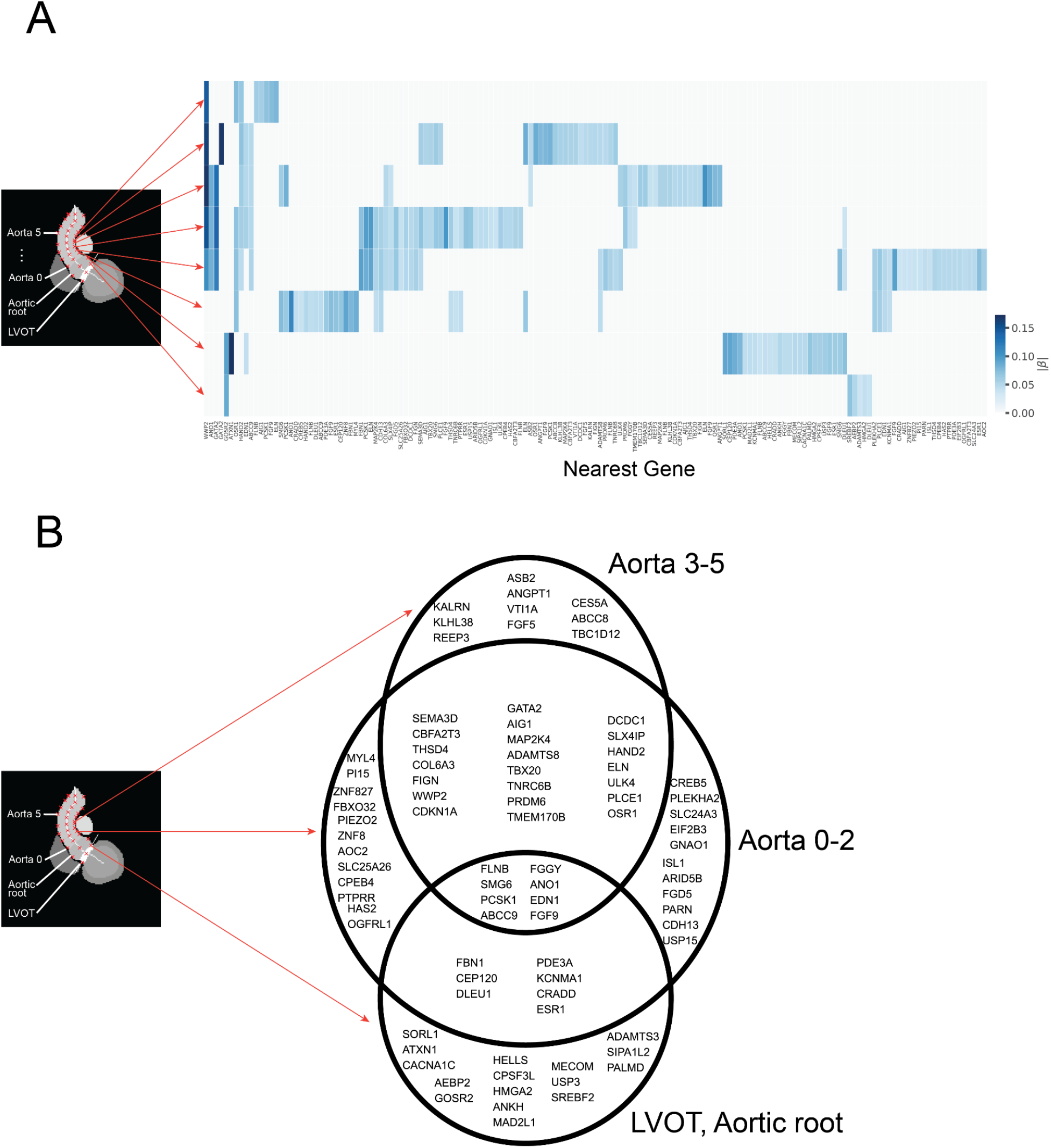
Clustering of significant loci **Panel A**: Row position denotes associated GWAS in fixed order corresponding to anatomical position as shown with red arrows. Columns denote SNPs associated with one or more traits at P < 5×10^−8^. Column labels display genes nearest to associated SNPs. In cases where multiple SNPs share a nearest gene, column labels display [nearest gene] _ [instance of gene’s occurrence]. Color indicates absolute effect size (|*β*|) per standard error of a SNP associated at P < 5×10^−8^. To illustrate anatomic relationship of genome-wide significant SNPs, heatmap column position is hierarchically clustered to group SNP associations of similar significance (P). For this illustrative purpose, SNPs not meeting genome-wide significance are assigned P = 0. **Panel B:** Venn diagram populated by genes nearest to loci found to be associated at P < 5×10^−8^ with one or more traits. Traits are binned into three groups based on proximity to the heart as labeled.

To investigate contributions from rare genetic variants to trait variation, we conducted rare variant association tests in 18,461 UK Biobank participants with both LVOT view imaging and exome sequencing data. No gene achieved Bonferroni significance in an exome-wide analysis.

### Genetic prediction of computed diameters are associated with thoracic aortic aneurysm

We next sought to investigate the relationship between the common-variant genetics underlying the diameters of the LVOT, aortic root, and ascending aortic tract and the incidence of aortic disease. Excluding participants whose data was used for the GWAS, we computed a polygenic risk score using the GWAS of each described phenotype using the autosomal, independently significant SNPs of the respective GWAS.

Using a Cox proportional hazards model adjusted for clinical risk factors, we analyzed the relationship between the resultant polygenic scores for individuals and incident thoracic aortic aneurysm, dissection, or rupture. We found that, for all eight diameters, polygenic scores corresponding to larger diameters positively correlated with incidence of disease at a significance level of P < 0.05. We observed the strongest effect size and significance for the polygenic score of Aorta 1 (the score representing a genetic prediction of the ascending aortic diameter approximately 13 mm distal to the aortic root, on a height-adjusted basis; n=427,016; HR=1.42 per standard deviation; CI=1.34-1.50, P=6.67×10^−21^). We further observed a gradient of signal in which both effect size and significance of the remaining seven scores was lesser in sequence with the associated diameter’s distance from Aorta 1 (**Figure 4A**). Finally, we observed that participants above the 90th percentile for a polygenic score of larger Aorta 1 diameter displayed a significantly increased incidence of thoracic aortic disease relative to the remaining participants in an unadjusted survival analysis (**Figure 4A**).

**Figure 4:**
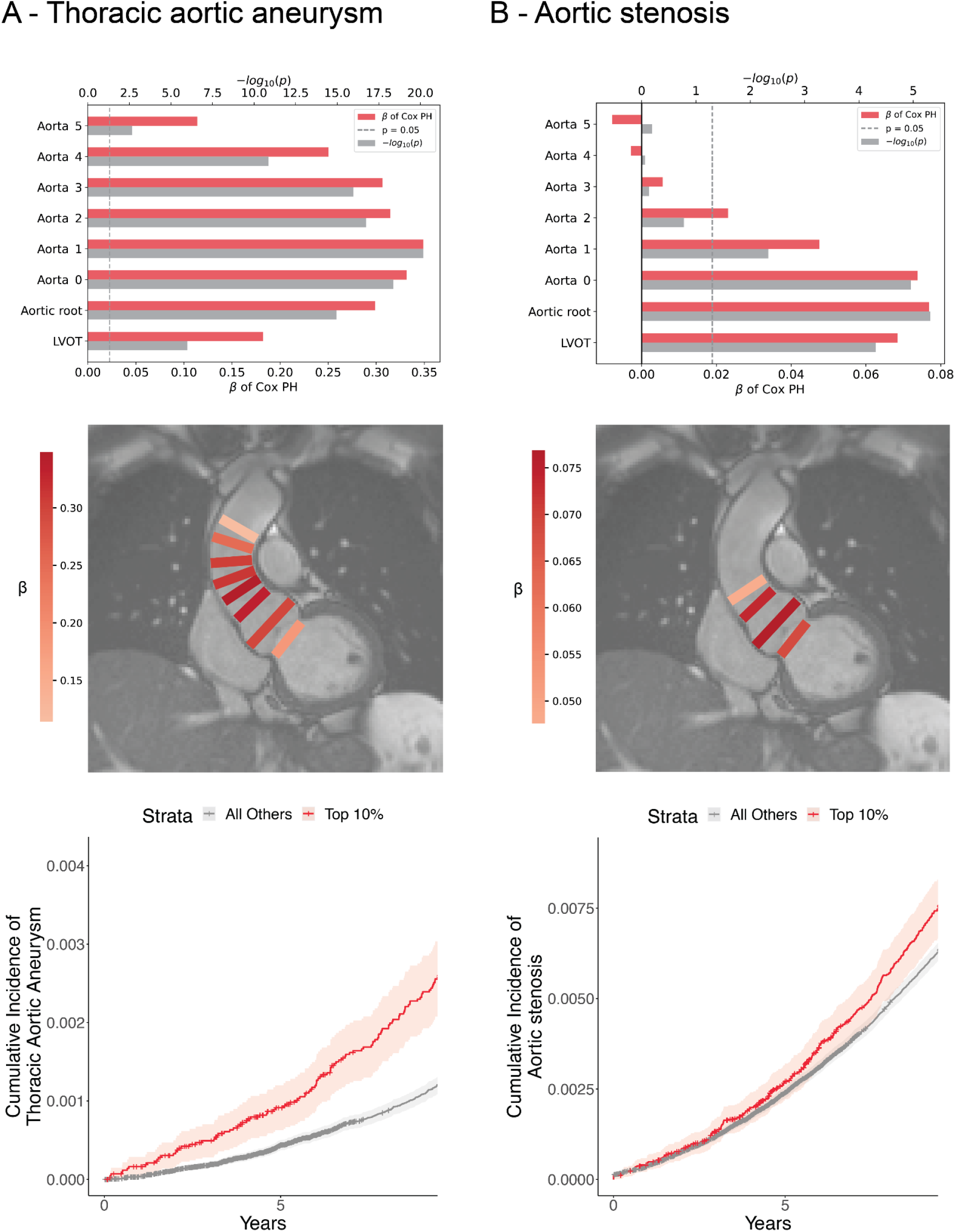
Disease prediction **Panel A:** Top: results of Cox proportional hazards models predicting incidence of thoracic aortic aneurysm using polygenic scores of given traits and clinical covariates. Higher polygenic scores correspond to larger trait diameters. Vertical position denotes trait used to create polygenic score in fixed, anatomic order. Horizontal bars denote effect size (*β*) and significance of models. Middle: Anatomic overlay of polygenic score performance. Rectangle color represents relative effect size (*β*); rectangle location represents approximate location of trait used to create polygenic score. Bottom: Kaplan-Meier plot denoting cumulative incidence of thoracic aortic aneurysm over time for strata of a polygenic score derived from Aorta 1. **Panel B:** Top: results of Cox proportional hazards models predicting incidence of aortic stenosis using polygenic scores of given traits and clinical covariates. Higher polygenic scores correspond to smaller trait diameters. Vertical position denotes trait used to create polygenic score in fixed, anatomic order. Horizontal bars denote effect size (*β*) and significance of models. Middle: Anatomic overlay of polygenic score performance. Rectangle color represents relative effect size (*β*); rectangle location represents approximate location of trait used to create polygenic score. Bottom: Kaplan-Meier plot denoting cumulative incidence of aortic stenosis over time for strata of a polygenic score derived from Aortic root.

### Genetic prediction of aortic root diameter is associated with aortic stenosis

We used a similar approach leveraging adjusted Cox proportional hazards models to investigate the relationship between the eight derived polygenic scores and incidence of aortic stenosis. In contrast to thoracic aortic aneurysm, which was positively associated with polygenic scores of larger diameters, we found that the scores corresponding to smaller values of the four most proximal diameters—LVOT, Aortic root, Aorta 0, and Aorta 1—significantly predicted incidence of aortic stenosis. We observed the strongest effect size and significance for the polygenic score of aortic root diameter, (n = 426,502, HR = 1.08 per standard deviation, CI 1.03-1.12, P = 5×10^−6^). Unlike for aortic aneurysm, where both proximal and distal polygenic scores were associated with disease, we observed that only aortic root, LVOT, and proximal aortic scores were associated with aortic stenosis: scores based on more distal diameters showed no significant correlation with incidence of disease. Participants above the 90th percentile of polygenic score derived from smaller Aortic root diameter displayed a significantly increased incidence of aortic stenosis relative to the remaining participants in an unadjusted survival analysis (**Figure 4B**).

## Discussion

We used deep learning to assess the size of the LVOT, aortic root, and ascending aorta using magnetic resonance imaging data in a large population-based biobank. To our knowledge this is the first GWAS of LVOT diameter and the first study to comparatively investigate the genetics underlying multiple diameters of the ascending aorta and their relative clinical significance. We identified 6 novel loci in the LVOT, 17 novel loci in the aortic root, and 23 novel loci in the ascending aorta, and assessed their association with thoracic aortic aneurysm and aortic stenosis. These findings permit several conclusions.

First, serial diameters of the LVOT-ascending aorta tract demonstrated distinct genetic underpinnings detectable even over short intervals of 10 millimeters or less. Where available, corroboration with prior knowledge suggests that these diverse genetic signatures are meaningful. For example, we found that a majority of loci significantly associated with ascending aortic diameter reproduce the results of our prior work investigating ascending aorta diameter via short axis imaging.^13^ We also observed genetic associations with aortic root diameter in agreement with prior echocardiography-driven work. For example, SNPs near *GOSR2* have previously been implicated in variations in aortic valve area, aortic root diameter, and other cardiac dimensions.^24–27^ Further, *PALMD* has been implicated as a susceptibility gene for calcific aortic stenosis.^24,28,29^ This agreement lends credence to interesting novel loci, such as aortic root diameter’s association with SNP rs80036911 on *MECOM*, a gene previously associated with left ventricular trabeculation.^26^ Developing a better understanding of the distinct drivers along the course of the LVOT-ascending aorta tract may ultimately enable more targeted therapies for subtypes of aortic pathology, such as root-predominant and root-sparing aortic aneurysm.

Second, we demonstrated spatial localization of a region of ascending aorta whose underlying genetics may be most relevant to incidence of aneurysm. Prediction was optimized with a polygenic score from Aorta 1 (a diameter about 13mm distal to the sinotubular junction) and signal sequentially diminished with subsequent diameters both proximally and distally. Loci found to be uniquely associated with this diameter include rs35296742 on *AOC2*, an amine oxidase, and rs1979974 on *ZNF827* which encodes a zinc finger protein observed to be involved in telomere synthesis and cell viability.^30,31^ Beyond individual targets, localizing the aorta diameter most salient for incidence of pathology may guide future efforts to develop imaging protocols to screen asymptomatic individuals at risk of thoracic aneurysm.

Third, we demonstrated that a polygenic score associated with a smaller aortic root diameter predicted incidence of aortic stenosis. Weaker predictive signals were seen for scores created for diameters of LVOT and proximal ascending aorta diameters, but we found no predictive signal for scores of more distal aorta diameters. Similar to the prediction of aneurysm, this finding implicates loci uniquely associated with aortic root diameter, or those uniquely associated with proximal diameters, as appealing targets for screening and therapeutics for aortic stenosis. One locus (lead SNP rs139939693, intronic to *USP3)* presents an interesting target for downstream research. *USP3*, encoding ubiquitin carboxyl-terminal hydrolase 13, has been previously implicated in atrial fibrillation, ECG morphology, and aortic root size (via echocardiography).^24,32,33^

Our work was subject to a number of limitations. All data were derived from deep learning models from cardiac magnetic resonance images. These models have imprecision that could be reduced with further training data. Like any deep learning model, these models can fail and produce non physiologic measurements when presented with images that contain features not seen in the training data. An advantage of the semantic segmentation approach in this work is that outliers can be visually inspected and the model re-trained as needed. The diameter measurements are one-dimensional estimates of three-dimensional structures and therefore cannot capture complete information about LVOT, aortic root, or ascending aorta size and morphology. With our quality control measures, we have attempted to ensure that the images selected for phenotype extraction display the LVOT and aorta near their maximal diameters, i.e. that the tract is not foreshortened. However, variations in anatomy, distance between diameters of interest, and the complex motion of the heart in the thorax introduce imprecision to this assumption. The deep learning models have not been tested outside of the specific devices and imaging protocols used by the UK Biobank and may not generalize to other data sets without additional fine-tuning. The study population is largely of European ancestry, similar to the remainder of UK Biobank, limiting generalizability of the findings to other populations. The individuals who underwent MRI in the UK Biobank tend to be healthier than the remainder of the UK Biobank population, which itself is healthier than the general population. Because we have used hospital-based diagnosis codes and procedural codes to identify individuals with disease our disease definitions are susceptible to misclassification. Finally, because echocardiography is the clinical standard for diagnosis of pathology of or near the aortic valve, it is possible that comparisons of polygenic scores’ ability to predict disease are biased in favor of scores derived from areas visible via echocardiography.

In summary, we used machine learning to obtain serial measurements of LVOT, aortic root, and ascending aorta diameter in a large population-based biobank. We found that the genetics underlying these measurements may be clinically relevant for the prediction of thoracic aortic aneurysm and aortic stenosis. For both aneurysm and stenosis, future work is warranted to determine whether a model incorporating a polygenic score and clinical risk factors might identify high-risk, asymptomatic individuals who would benefit from screening via thoracic imaging.

### Sources of funding

This work was supported by the Fondation Leducq (14CVD01), and by grants from the National Institutes of Health to Dr. Pirruccello (K08HL159346), Dr. Ellinor (1RO1HL092577, K24HL105780) and Dr. Ho (R01HL134893, R01HL140224, K24HL153669). This work was also supported by a Sarnoff Cardiovascular Research Foundation Scholar Award to Dr. Pirruccello. Dr. Nauffal is supported by NIH grant 5T32HL007604-35. Dr. Lubitz is supported by NIH grant R01HL139731 and American Heart Association 18SFRN34250007. This work was supported by a grant from the American Heart Association Strategically Focused Research Networks to Dr. Ellinor. Dr. Lindsay is supported by the Fredman Fellowship for Aortic Disease and the Toomey Fund for Aortic Dissection Research. This work was funded by a collaboration between the Broad Institute and IBM Research.

### Disclosures

Dr. Pirruccello has served as a consultant for Maze Therapeutics. Dr. Lubitz receives sponsored research support from Bristol Myers Squibb / Pfizer, Bayer AG, Boehringer Ingelheim, and Fitbit, and has consulted for Bristol Myers Squibb / Pfizer and Bayer AG, and participates in a research collaboration with IBM. Dr. Ng is employed by IBM Research. Dr. Batra receives sponsored research support from Bayer AG and IBM, and has consulted for Novartis and Prometheus Biosciences. Dr. Ho has received research grant support from Bayer AG focused on machine-learning and cardiovascular disease, and research supplies from EcoNugenics. Dr. Philippakis is supported by a grant from Bayer AG to the Broad Institute focused on machine learning for clinical trial design. Dr. Ellinor is supported by a grant from Bayer AG to the Broad Institute focused on the genetics and therapeutics of cardiovascular diseases. Dr. Ellinor has also served on advisory boards or consulted for Bayer AG, Quest Diagnostics, MyoKardia and Novartis. Remaining authors report no disclosures.

## Methods

### Study design, IRB and ethical considerations

The UK Biobank is a phenotyped, prospective, population-based cohort of 500,000 individuals between the ages of 40-69 recruited by mail from 2006-2010.^34^ Within the UK Biobank, some 100,000 individuals underwent cardiac magnetic resonance (CMR) imaging.^35,36^ Cardiac magnetic resonance imaging was performed with 1.5 Tesla scanners (MAGNETOM Aera, Siemens Healthcare), using electrocardiographic gating for cardiac synchronization.^36^ In total, quality control measures detailed in our previous work suggest that in this study, 2.3 million images from 45,000 individuals would be eligible for analysis.^13^ UK Biobank access is provided under application 7089. Analysis is approved by the Mass General Brigham institutional review board (protocol 2003P001563). Identifiers of UK Biobank subjects who have withdrawn consent since the time of their enrollment are regularly distributed by the repository; these subjects are excluded from future analyses.

### Annotation

Segmentation maps of the cine images have been traced manually (MN) as supervised by a cardiologist (JPP). In our prior work we noted that a similar downstream deep learning model for CMR images produced optimum downstream pixel accuracy when trained with at least 116 manually annotated images, and that additional accuracy gleaned from training with up to 500 manually annotated images was modest.^13^ Thus, in the present work 250 samples were chosen at random, manually segmented, and then used to train a deep learning model with fastai v1.0.5945.^18^ The model was U-net derived with an encoder pre-trained on ImageNet to optimize for the recognition of natural shapes.^19,21^ Training and validation subsets comprised 80% and 20% of the samples, respectively. The training model was found to achieve 99.8% pixel segmentation accuracy (Jaccard agreement 84.3%) for annotation of the LVOT in the held-out validation set. Pulmonary artery segmentation was used as a secondary annotation metric; for this structure the pixel segmentation accuracy was found to be 99.9% (Jaccard agreement 87.3%).

### Calculation of diameters of anatomic structures

The LVOT, the aortic root, and the ascending aorta, are captured in the long axis of the contiguous tract they form. As annotated in this plane, the LVOT, aortic root, and proximal part of the ascending aorta often appear to be rectangular. To extract their diameters, first, the centerline of the contiguous tract formed by the LV cavity, LVOT, aortic root, and ascending aorta was calculated using the algorithm skeletonize implemented by the image processing package scikit-image.^22,23^ Orthogonal measurements to the tract centerline were taken at the respective centroids of the LVOT and aortic root complex. These normal lines were then intersected with the boundaries of the respective structure to give two coordinates representing the bounds of the diameter measurements. The Euclidean distance between the diameter boundaries, converted from pixels to millimeters based on image metadata, represented the diameters of the LVOT and aortic root (**Figure 1**).

For the ascending aorta, rather than a single diameter taken at the centroid, we took several diameters that we sought to align between participants to represent similar levels of aorta. To achieve this, we used height as a proxy for aorta length. We defined the centerline at the border with the sinotubular junction as a reference point. We then identified the point on the centerline 3 mm distal to this border, and via a perpendicular line as for the proximal structures, measured a diameter at this point (“Aorta 0”). We then took subsequent diameters of the aorta at intervals of 10 mm multiplied by the ratio of the participant’s height to the mean height of the dataset (i.e. 169 cm). For example, a participant of mean height would have diameters of the aorta taken, starting at the border with the aortic root, at 3 mm, 13 mm, 23 mm, and so on (see **Supplemental Table 5** for descriptions of all phenotypes). These measurements are taken closer together for a shorter-than-average participant and farther apart for a taller-than-average participant. Because a variable length of aorta was visualized across participants (and across studies) because of variation in its orientation relative to the imaging plane, the number of ascending aorta diameters that we could measure for each participant was, similarly, variable.

### Quality control of image segmentation and phenotype extraction

Images were filtered to those containing one component for each of: left ventricle cavity, LVOT, and aortic root. Images containing an LV cavity of less than 100 pixels were discarded due to biologic implausibility. Phenotypic measurements of LVOT diameter, aortic root diameter, or ascending aorta diameter at any level less than 1cm were discarded due to biologic implausibility. If a given participant was imaged on multiple occasions, we kept only the first study in order to facilitate comparisons with prior published work. It was observed that greater than 50% of the participants in the dataset had measurements of the aorta at levels Aorta 0 through Aorta 5. More distal levels were thus excluded due to incompleteness.

We observed that the contiguous tract of LVOT, aortic root, and ascending aorta were most often captured in correct planar arrangement at the beginning of the MRI CINE: a point of the cardiac cycle corresponding to left ventricular end-diastole. Other points of the cardiac cycle, for example ventricular end-systole, failed to capture the aorta in-plane. Therefore, for a given MRI study, we chose the first frame for subsequent analysis, or the second frame if the first frame had failed upstream QC. We confirmed that the mean LV cavity size using this approach very closely approximated the corresponding value if instead the maximum LV cavity for each participant were chosen. We thus concluded that choosing the first or second frame of the study appropriately maximized the likelihood of a correct planar arrangement while remaining very close to true ventricular end-diastole.

To identify any remaining images with an incorrect planar arrangement, we exploited the fact that we have previously measured the diameter of the ascending aorta in its cross section (short axis) using a different deep learning model trained by different operators.^13^ We compared the present work’s estimates of structure diameters against the previously published diameters of ascending aorta in its short axis. We found that the LVOT-view-derived diameters for ascending aorta at position Aorta 2 correlated most strongly with the previously published phenotype (Pearson’s r = 0.83). We then investigated the ratio of LVOT-view-derived diameters to the previously published short axis diameter and found that the ratio closely approached parity at aorta position 2 (median = 0.98, interquartile range = 0.08). As such, we hypothesized that the previously investigated short axis plane was likely taken most often nearest to the position labeled Aorta 2 in our present study. As discussed in **Methods: Calculation of diameters of anatomic structures**, this point corresponds to approximately 23 mm distal to the border with the aortic root for a participant of average height.

We further hypothesized that points for which the relationship between the LVOT-view-derived diameter Aorta 2 and the short axis diameter of the aorta diverges significantly from a linear relationship represent out-of-plane images. We thus constructed a linear model relating these two phenotypes and obtained 99% prediction intervals for all values of the independent variable: ascending aorta taken from the LVOT view at position 2. Those points with a corresponding ascending aorta diameter derived from its short axis view fell outside the 99% prediction intervals were discarded. We excluded participants with prevalent atrial fibrillation, heart failure, myocardial infarction, or coronary artery disease prior to MRI. We finally excluded participants with lack of genetic data, sample QC missing rate greater than or equal to 2%, sex chromosome aneuyploidy, or outlier status for heterozygosity (**Supplemental Figure 1**). After final quality control, up to 33,870 participants remained to be included in a GWAS; sample size decreased for GWAS traits of more distal ascending aorta given its variable visualization (**Supplemental Table 3**).

### Genotyping and genome-wide association studies

Genotype information was obtained using either the UK BiLEVE or UK Biobank Axiom arrays; they were then imputed into the Haplotype Reference Consortium panel and the UK10K+1000 Genomes panel.^37^ Participants were filtered to exclude those without imputed genetic data and those with a genotypic call rate < 0.98. Heritability values of structures of interest were then calculated using BOLT-REML on the genotyped variants in the UK Biobank.^38^ Covariates included age at time of MRI, the first five principal components of ancestry, sex, the genotyping array, and the MRI scanner’s unique identifier.^39^

Genome-wide association was tested using REGENIE 2.0.2, using the full autosomal panel of directly genotyped SNPs as the genetic relationship matrix and structure diameters as continuous phenotypes, as discussed.^40^ GWAS covariates included age at time of MRI, the first five principal components of ancestry, sex, the genotyping array, and the MRI scanner’s unique identifier.^39^ The genome-wide significance threshold for SNPs was P < 5×10^−8^. To identify independently associated variants, linkage disequilibrium clumping was performed in the same participants used to create the GWAS using plink-1.9 using a 5 megabase radius, an r^2^ cutoff of 0.001, and the genome-wide P threshold.^41^ For comparison of loci reaching genome-wide significance for multiple traits, a given locus was assigned a shared “global” locus identifying number with any other loci within 500 kilobases. Lead SNPs were tested for deviation from Hardy-Weinberg equilibrium (HWE) at the commonly-used threshold P < 1×10^−6^. BOLT-REML v2.3.4 was used to assess the SNP heritability of all phenotypes.^38^

### Exome sequencing in UK Biobank

We conducted an exome sequencing analysis on over 200,000 exomes released by the UK Biobank. Samples from the UK Biobank were chosen for exome sequencing based on enrichment for MRI data and linked health records.^42^ Exome sequencing was performed by Regeneron and reprocessed centrally by the UK Biobank following the Functional Equivalent pipeline.^43^ Exomes were captured with the IDT xGen Exome Research Panel v1.0.The basic design targets 39Mbp of the human genome (19,396 genes). Multiplexed samples were sequenced with dual-indexed 75×75bp paired-end reads on the Illumina NovaSeq 6000 platform using S2 (first 50k samples) and S4 flow cells (subsequent 150k samples). Alignment to GRCh38 was performed in an alt-aware manner as described in the Functional Equivalence protocol. Individual level VCF files were combined and joint-genotyped using GLnexus after Variants were called per-sample using DeepVariant. We applied hard filters low-quality genotypes:

1. For homozygous reference calls: Genotype Quality < 20; Genotype Depth < 10; Genotype Depth > 200
2. For heterozygous calls: (A1 Depth + A2 Depth)/Total Depth < 0.9; A2 Depth/Total Depth < 0.2; Genotype likelihood[ref/ref] < 20; Genotype Depth < 10; Genotype Depth > 200
3. For homozygous alternative calls: (A1 Depth + A2 Depth)/Total Depth < 0.9; A2 Depth/Total Depth < 0.9; Genotype likelihood[ref/ref] < 20; Genotype Depth < 10; Genotype Depth > 200

Then, we performed variant level filters. Low-quality variants were removed followed by: Call rate < 90%, Hardy-Weinberg Equilibrium p-value < 10-15, present in Ensembel low-complexity regions, monomorphic variants in the final dataset. Finally, we calculated a few sample quality metrics and removed low-quality samples. Samples with withdrawn consent, sex mismatch, call rate < 90%, outliers (8 standard deviations apart from mean) of transition/transversion ratio, heterozygote/homozygote ratio, SNP/Indel ratio, and the number of singletons were removed.

Variants were annotated with the Ensembl Variant Effect Predictor version 95 using the --pick-allele flag.^44^ LOFTEE was used to identify high-confidence loss of function variants: stop-gain, splice-site disrupting, and frameshift variants.^45^ To understand population-level frequencies of variants, we annotated 5 continental variant frequencies using gnomAD v2.

### Rare variant association test

We conducted a collapsing burden test to assess the impact of loss-of-function variants in up to 18,461 participants who had measurements of diameters of interest and exome sequencing data available. Variants with MAF ≥ 0.001 or continental maximum MAF ≥ 0.001 were excluded. Using the LOFTEE “high-confidence” loss-of-function variants without a flag, we tested whether loss-of-function carrier status was associated with ascending aorta traits using the linear mixed-effect approach implemented in the R-package GENESIS. The model was adjusted for age at MRI, sex, the MRI serial number, sequencing batch, associated PCs, and empirical kinship matrix. Protein-coding genes with the number of carriers <10 were removed from the results.

### Comparison with prior work

We sought to compare our GWAS results with those of comparable studies. We identified one that investigated the genetics of ascending aorta diameter at a comparable anatomic location and one that investigated the genetics of aortic root diameter at a comparable anatomic location.^13,24^ To our knowledge, no prior study investigated the genetics of the LVOT diameter in a comparable way. For both aorta and aortic root, we deemed a trait novel if it was both (1) outside 500 kilobases from all SNPs in the prior work and (2) not in linkage disequilibrium with any SNP in the prior work. To query linkage disequilibrium we queried all SNPs with rsIDs against the associated prior work’s SNPs using LDlink, specifying European populations to best match the UK Biobank participants and using an LD cutoff of r^2^ > 0.01.^46^ In one instance, LDlink was unable to resolve a SNP described in the present work (at chr5:15005465, associated with aortic root diameter). In this case we manually confirmed that there were no previously described SNPs in the associated prior work within 100 megabases.

### Trait-disease correlation

We sought to validate our diameters by investigating their correlations to prevalent disease (**Supplemental Table 6**). To do so we constructed Cox proportional hazards models investigating the presence or absence of a disease of interest as predicted by the inverse normal diameter of interest as well as covariates including age at time of MRI, sex, MRI serial number, genotyping array, and the five first principal components of ancestry.

### Aortic disease codes

International Classification of Diseases version 10 (ICD-10) codes and Office of Population Censuses and Surveys Classification of Interventions and Procedures version 4 (OPCS-4) codes used to define diseases of interest are detailed in Supplementary Table 1. These definitions were used for GWAS participant exclusion and polygenic score assessment.

### Association between polygenic risk scores and incident disease

Using PRScs (https://github.com/getian107/PRScs; git hash 43128be7fc9ca16ad8b85d8754c538bcfb7ec7b4), we computed polygenic risk scores for participants in the UK Biobank with genetic data available.^47^ We computed a separate score using the GWAS of each described phenotype, using the autosomal, independently significant SNPs of the respective GWAS. We excluded participants whose data was used for the GWAS.

We analyzed the relationship between the resultant polygenic scores for individuals and several events using Cox proportional hazards models that were also adjusted for clinical risk factors. The events included incident thoracic aortic aneurysm (427,016 individuals, 743 events) and aortic stenosis (426,502 individuals, 3,604 events). There are limited data regarding clinical risk factors for thoracic aortic aneurysm outside of associated syndromes and family history, so we chose putatively relevant covariates based in part on inference from evidence in the abdominal aortic aneurysm literature.^48^ These covariates included sex, body mass (the cubic natural spline of BMI), age (the cubic natural spline of age at enrollment), and blood pressure (the cubic natural splines of systolic and diastolic blood pressure measurements). We also adjusted for other covariates including the cubic natural spline of height, the cubic natural spline of weight, the genotyping array, and the first five principal components of ancestry. This analysis was performed separately for each of the polygenic risk scores listed above.

## Data Availability

All data produced in the present study are available upon reasonable request to the authors

## References

1. Orts-Llorca F, Puerta Fonolla J, Sobrado J. The formation, septation and fate of the truncus arteriosus in man. Journal of anatomy. 1982;134(Pt 1):41–56.

2. Grotenhuis HB, Dallaire F, Verpalen IM, van den Akker MJE, Mertens L, Friedberg MK. Aortic Root Dilatation and Aortic-Related Complications in Children After Tetralogy of Fallot Repair. Circulation. Cardiovascular imaging. 2018;11(12):e007611.

3. Axt-Fliedner R, Kreiselmaier P, Schwarze A, Krapp M, Gembruch U. Development of hypoplastic left heart syndrome after diagnosis of aortic stenosis in the first trimester by early echocardiography. Ultrasound in obstetrics & gynecology: the official journal of the International Society of Ultrasound in Obstetrics and Gynecology. 2006;28(1):106–109.

4. Howard DPJ, Banerjee A, Fairhead JF, Perkins J, Silver LE, Rothwell PM, Oxford Vascular Study. Population-based study of incidence and outcome of acute aortic dissection and premorbid risk factor control: 10-year results from the Oxford Vascular Study. Circulation. 2013;127(20):2031–2037.

5. Renard M, Francis C, Ghosh R, Scott AF, Witmer PD, Adès LC, Andelfinger GU, Arnaud P, Boileau C, Callewaert BL, Guo D, Hanna N, Lindsay ME, Morisaki H, Morisaki T, et al. Clinical Validity of Genes for Heritable Thoracic Aortic Aneurysm and Dissection. Journal of the American College of Cardiology. 2018;72(6):605–615.

6. Fann JI. Descending thoracic and thoracoabdominal aortic aneurysms. Coronary artery disease. 2002;13(2):93–102.

7. Guo D-C, Papke CL, He R, Milewicz DM. Pathogenesis of thoracic and abdominal aortic aneurysms. Annals of the New York Academy of Sciences. 2006;1085:339–352.

8. Vapnik JS, Kim JB, Isselbacher EM, Ghoshhajra BB, Cheng Y, Sundt TM 3rd, MacGillivray TE, Cambria RP, Lindsay ME. Characteristics and Outcomes of Ascending Versus Descending Thoracic Aortic Aneurysms. The American journal of cardiology. 2016;117(10):1683–1690.

9. Jondeau G, Boileau C. Familial thoracic aortic aneurysms. Current opinion in cardiology. 2014;29(6):492–498.

10. Verstraeten A, Luyckx I, Loeys B. Aetiology and management of hereditary aortopathy. Nature reviews. Cardiology. 2017;14(4):197–208.

11. Pinard A, Jones GT, Milewicz DM. Genetics of thoracic and abdominal aortic diseases. Circulation research. 2019;124(4):588–606.

12. Ashvetiya T, Fan SX, Chen Y-J, Williams CH, O’Connell JR, Perry JA, Hong CC. Identification of novel genetic susceptibility loci for thoracic and abdominal aortic aneurysms via genome-wide association study using the UK Biobank Cohort. PloS one. 2021;16(9):e0247287.

13. Pirruccello JP, Chaffin MD, Chou EL, Fleming SJ, Lin H, Nekoui M, Khurshid S, Friedman SN, Bick AG, Arduini A, Weng L-C, Choi SH, Akkad A-D, Batra P, Tucker NR, et al. Deep learning enables genetic analysis of the human thoracic aorta. Nature Genetics. Accepted.

14. Murillo H, Lane MJ, Punn R, Fleischmann D, Restrepo CS. Imaging of the aorta: embryology and anatomy. Seminars in ultrasound, CT, and MR. 2012;33(3):169–190.

15. Verzi MP, McCulley DJ, De Val S, Dodou E, Black BL. The right ventricle, outflow tract, and ventricular septum comprise a restricted expression domain within the secondary/anterior heart field. Developmental biology. 2005;287(1):134–145.

16. Waldo KL, Hutson MR, Ward CC, Zdanowicz M, Stadt HA, Kumiski D, Abu-Issa R, Kirby ML. Secondary heart field contributes myocardium and smooth muscle to the arterial pole of the developing heart. Developmental biology. 2005;281(1):78–90.

17. Jiang X, Rowitch DH, Soriano P, McMahon AP, Sucov HM. Fate of the mammalian cardiac neural crest. Development. 2000;127(8):1607–1616.

18. Howard J, Gugger S. Fastai: A layered API for deep learning. Information. An International Interdisciplinary Journal. 2020;11(2):108.

19. Ronneberger O, Fischer P, Brox T. U-net: Convolutional networks for biomedical image segmentation. In: International Conference on Medical image computing and computer-assisted intervention. Springer; 2015:234–241.

20. Russakovsky O, Deng J, Su H, Krause J, Satheesh S, Ma S, Huang Z, Karpathy A, Khosla A, Bernstein M, Berg AC, Fei-Fei L. ImageNet Large Scale Visual Recognition Challenge. International journal of computer vision. 2015;115(3):211–252.

21. Deng J, Dong W, Socher R, Li L-J, Li K, Fei-Fei L. Imagenet: A large-scale hierarchical image database. In: 2009 IEEE conference on computer vision and pattern recognition. Ieee; 2009:248–255.

22. Zhang TY, Suen CY. A fast parallel algorithm for thinning digital patterns. Communications of the ACM. 1984;27(3):236–239.

23. van der Walt S, Schönberger JL, Nunez-Iglesias J, Boulogne F, Warner JD, Yager N, Gouillart E, Yu T, scikit-image contributors. scikit-image: image processing in Python. PeerJ. 2014;2:e453.

24. Wild PS, Felix JF, Schillert A, Teumer A, Chen M-H, Leening MJG, Völker U, Großmann V, Brody JA, Irvin MR, Shah SJ, Pramana S, Lieb W, Schmidt R, Stanton AV, et al. Large-scale genome-wide analysis identifies genetic variants associated with cardiac structure and function. The Journal of clinical investigation. 2017;127(5):1798–1812.

25. Córdova-Palomera A, Tcheandjieu C, Fries JA, Varma P, Chen VS, Fiterau M, Xiao K, Tejeda H, Keavney BD, Cordell HJ, Tanigawa Y, Venkataraman G, Rivas MA, Ré C, Ashley E, et al. Cardiac Imaging of Aortic Valve Area From 34 287 UK Biobank Participants Reveals Novel Genetic Associations and Shared Genetic Comorbidity With Multiple Disease Phenotypes. Circulation. Genomic and precision medicine. 2020;13(6):e003014.

26. Meyer HV, Dawes TJW, Serrani M, Bai W, Tokarczuk P, Cai J, de Marvao A, Henry A, Lumbers RT, Gierten J, Thumberger T, Wittbrodt J, Ware JS, Rueckert D, Matthews PM, et al. Genetic and functional insights into the fractal structure of the heart. Nature. 2020;584(7822):589–594.

27. Yu M, Tcheandjieu C, Georges A, Xiao K, Tejeda H, Dina C, Le Tourneau T, Fiterau I, Judy R, Tsao N, Amgalan D, Munger CJ, Engreitz JM, Damrauer S, Bouatia-Naji N, et al. Computational estimates of mitral annular diameter in systole and diastole cardiac cycle reveal novel genetic determinants of valve function and disease. bioRxiv. 2020.

28. Thériault S, Gaudreault N, Lamontagne M, Rosa M, Boulanger M-C, Messika-Zeitoun D, Clavel M-A, Capoulade R, Dagenais F, Pibarot P, Mathieu P, Bossé Y. A transcriptome-wide association study identifies PALMD as a susceptibility gene for calcific aortic valve stenosis. Nature communications. 2018;9(1):988.

29. Li Z, Gaudreault N, Arsenault BJ, Mathieu P, Bossé Y, Thériault S. Phenome-wide analyses establish a specific association between aortic valve PALMD expression and calcific aortic valve stenosis. Communications biology. 2020;3(1):477.

30. Kaitaniemi S, Elovaara H, Grön K, Kidron H, Liukkonen J, Salminen T, Salmi M, Jalkanen S, Elima K. The unique substrate specificity of human AOC2, a semicarbazide-sensitive amine oxidase. Cellular and molecular life sciences: CMLS. 2009;66(16):2743–2757.

31. Conomos D, Reddel RR, Pickett HA. NuRD–ZNF827 recruitment to telomeres creates a molecular scaffold for homologous recombination. Nature structural & molecular biology. 2014;21(9):760–770.

32. Roselli C, Chaffin MD, Weng L-C, Aeschbacher S, Ahlberg G, Albert CM, Almgren P, Alonso A, Anderson CD, Aragam KG, Arking DE, Barnard J, Bartz TM, Benjamin EJ, Bihlmeyer NA, et al. Multi-ethnic genome-wide association study for atrial fibrillation. Nature genetics. 2018;50(9):1225–1233.

33. Verweij N, Benjamins J-W, Morley MP, van de Vegte YJ, Teumer A, Trenkwalder T, Reinhard W, Cappola TP, van der Harst P. The Genetic Makeup of the Electrocardiogram. Cell systems. 2020;11(3):229–238.e5.

34. Sudlow C, Gallacher J, Allen N, Beral V, Burton P, Danesh J, Downey P, Elliott P, Green J, Landray M, Others. UK biobank: an open access resource for identifying the causes of a wide range of complex diseases of middle and old age. PLoS medicine. 2015;12(3):e1001779.

35. Petersen SE, Matthews PM, Bamberg F, Bluemke DA, Francis JM, Friedrich MG, Leeson P, Nagel E, Plein S, Rademakers FE, Others. Imaging in population science: cardiovascular magnetic resonance in 100,000 participants of UK Biobank-rationale, challenges and approaches. Journal of cardiovascular magnetic resonance: official journal of the Society for Cardiovascular Magnetic Resonance. 2013;15(1):46.

36. Petersen SE, Matthews PM, Francis JM, Robson MD, Zemrak F, Boubertakh R, Young AA, Hudson S, Weale P, Garratt S, Others. UK Biobank’s cardiovascular magnetic resonance protocol. Journal of cardiovascular magnetic resonance: official journal of the Society for Cardiovascular Magnetic Resonance. 2015;18(1):8.

37. Bycroft C, Freeman C, Petkova D, Band G, Elliott LT, Sharp K, Motyer A, Vukcevic D, Delaneau O, O’Connell J, Others. The UK Biobank resource with deep phenotyping and genomic data. Nature. 2018;562(7726):203–209.

38. Loh P-R, Tucker G, Bulik-Sullivan BK, Vilhjálmsson BJ, Finucane HK, Salem RM, Chasman DI, Ridker PM, Neale BM, Berger B, Others. Efficient Bayesian mixed-model analysis increases association power in large cohorts. Nature genetics. 2015;47(3):284.

39. Eveborn GW, Schirmer H, Heggelund G, Lunde P, Rasmussen K. The evolving epidemiology of valvular aortic stenosis. the Tromsø study. Heart. 2013;99(6):396–400.

40. Mbatchou J, Barnard L, Backman J, Marcketta A, Kosmicki JA, Ziyatdinov A, Benner C, O’Dushlaine C, Barber M, Boutkov B, Habegger L, Ferreira M, Baras A, Reid J, Abecasis G, et al. Computationally efficient whole-genome regression for quantitative and binary traits. Nature genetics. 2021;53(7):1097–1103.

41. Chang CC, Chow CC, Tellier LC, Vattikuti S, Purcell SM, Lee JJ. Second-generation PLINK: rising to the challenge of larger and richer datasets. GigaScience. 2015;4(1):s13742–015.

42. Van Hout CV, Tachmazidou I, Backman JD, Hoffman JD, Liu D, Pandey AK, Gonzaga-Jauregui C, Khalid S, Ye B, Banerjee N, Li AH, O’Dushlaine C, Marcketta A, Staples J, Schurmann C, et al. Exome sequencing and characterization of 49,960 individuals in the UK Biobank. Nature. 2020;586(7831):749–756.

43. Regier AA, Farjoun Y, Larson DE, Krasheninina O, Kang HM, Howrigan DP, Chen B-J, Kher M, Banks E, Ames DC, English AC, Li H, Xing J, Zhang Y, Matise T, et al. Functional equivalence of genome sequencing analysis pipelines enables harmonized variant calling across human genetics projects. Nature communications. 2018;9(1):4038.

44. McLaren W, Gil L, Hunt SE, Riat HS, Ritchie GRS, Thormann A, Flicek P, Cunningham F. The Ensembl Variant Effect Predictor. Genome biology. 2016;17(1):122.

45. Karczewski KJ, Francioli LC, Tiao G, Cummings BB, Alföldi J, Wang Q, Collins RL, Laricchia KM, Ganna A, Birnbaum DP, Gauthier LD, Brand H, Solomonson M, Watts NA, Rhodes D, et al. Author Correction: The mutational constraint spectrum quantified from variation in 141,456 humans. Nature. 2021;590(7846):E53.

46. Machiela MJ, Chanock SJ. LDassoc: an online tool for interactively exploring genome-wide association study results and prioritizing variants for functional investigation. Bioinformatics. 2018;34(5):887–889.

47. Ge T, Chen C-Y, Ni Y, Feng Y-CA, Smoller JW. Polygenic prediction via Bayesian regression and continuous shrinkage priors. Nature communications. 2019;10(1):1776.

48. Kent KC, Zwolak RM, Egorova NN, Riles TS, Manganaro A, Moskowitz AJ, Gelijns AC, Greco G. Analysis of risk factors for abdominal aortic aneurysm in a cohort of more than 3 million individuals. Journal of vascular surgery. 2010;52(3):539–548.

