## Supplemental Figures for "Using Machine Learning to Elucidate the Spatial and Genetic Complexity of the Ascending Aorta"

**Supplemental Figure 1:** Quality control for GWAS

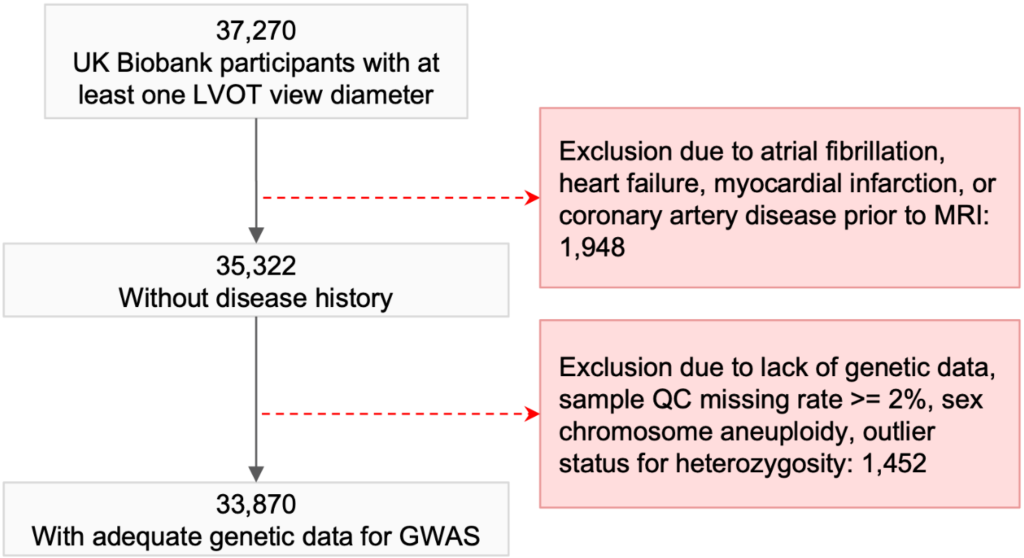

**Supplemental Figure 2:** Distributions of extracted diameters

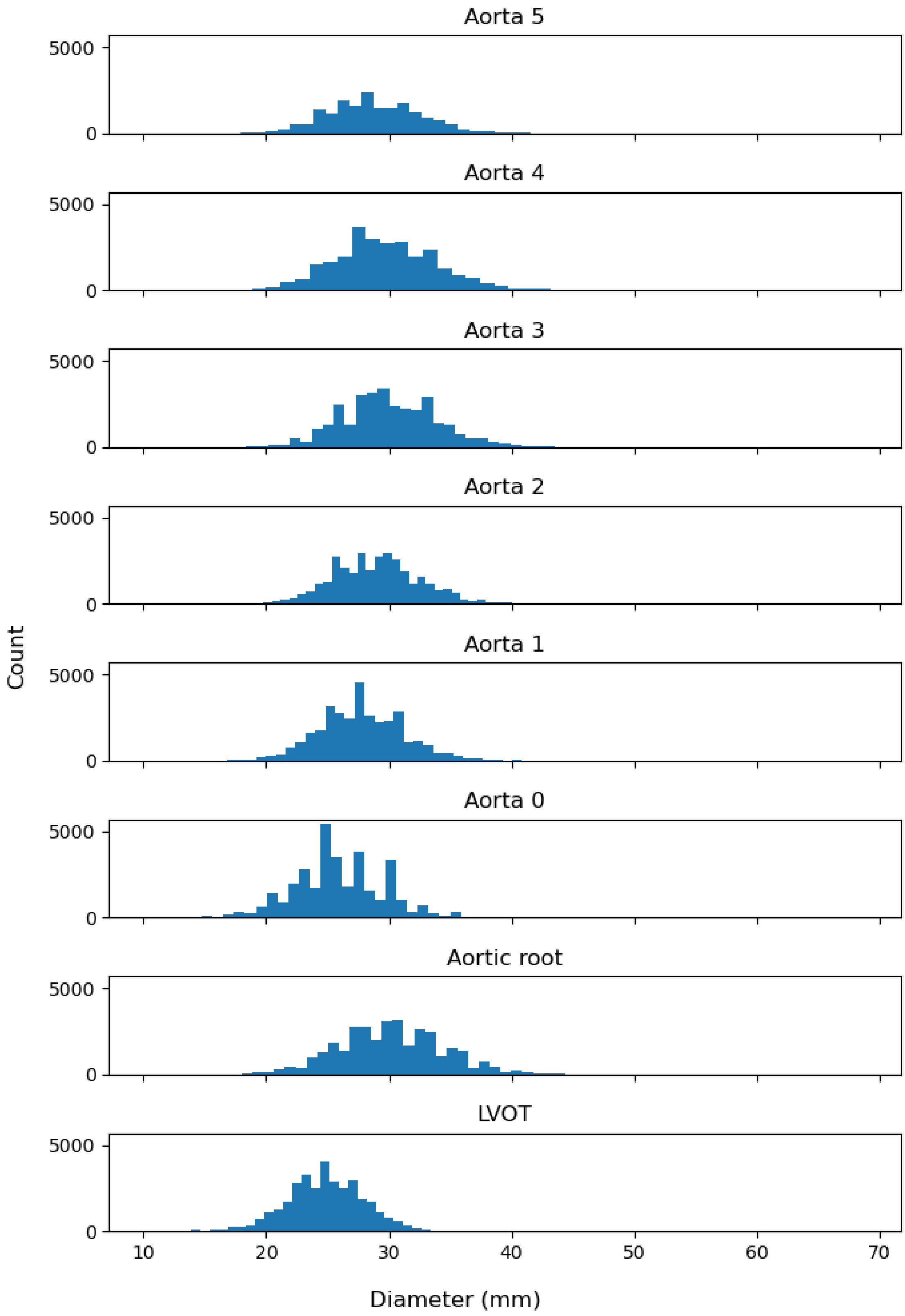

**Supplemental Figure 3:** Genetic correlations between traits

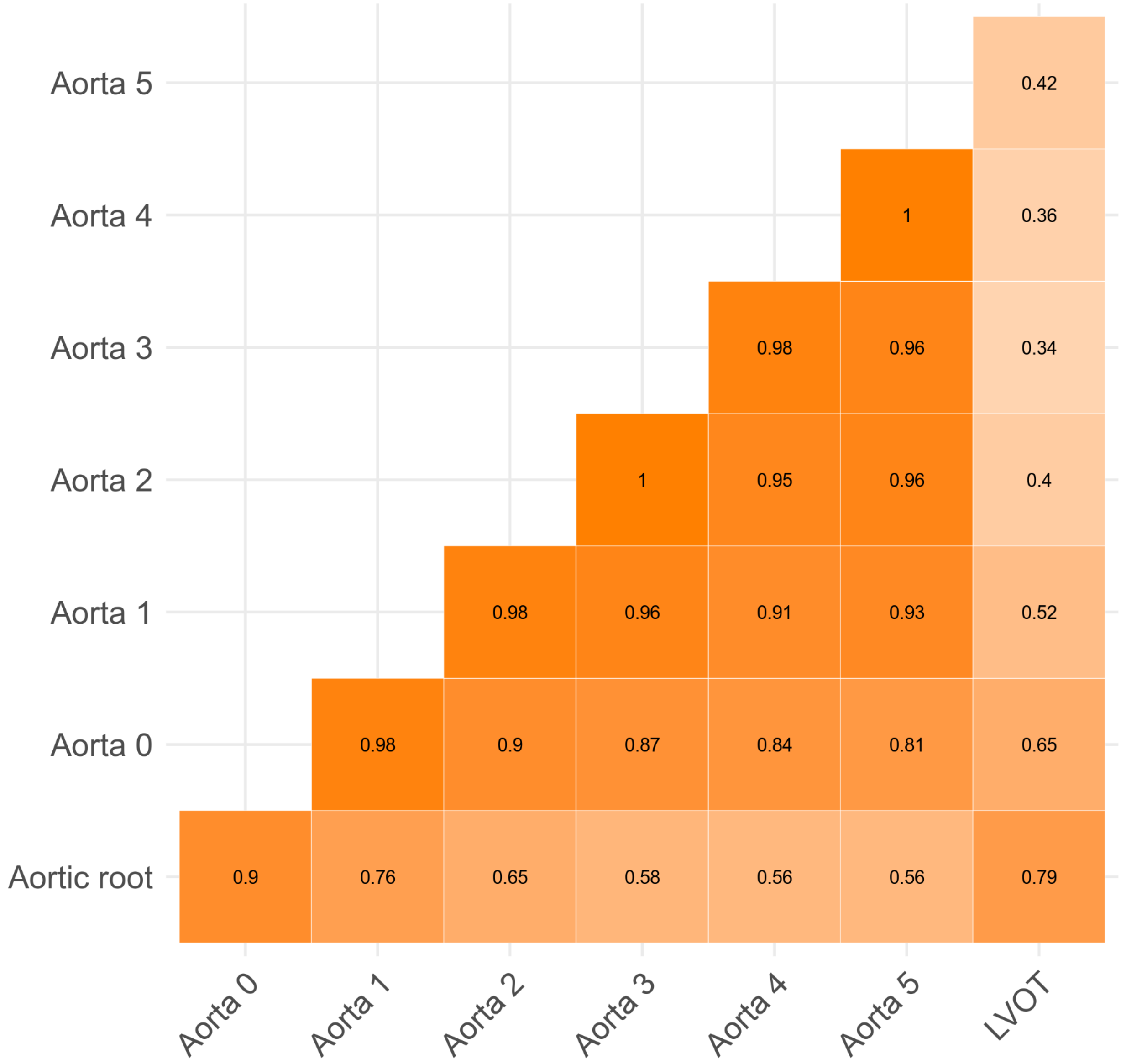
